# Modeling COVID-19 dynamics in the sixteen West African countries

**DOI:** 10.1101/2020.09.04.20188532

**Authors:** Sewanou H. Honfo, Hémaho B. Taboe, Romain Glèlè Kakaï

**Affiliations:** Laboratoire de Biomathématiques et d’Estimations Forestières, Université d’Abomey-Calavi, Benin

**Keywords:** COVID-19, detection rate, peak time and size, final epidemic size, attack ratio\, Africa

## Abstract

The COVID-19 pandemic is currently causing several damages to the world, especially in the public health sector. Due to identifiability problems in parameters’ estimation of complex compartmental models, this study considered a simple deterministic susceptible–infectious–recovered (SIR)-type model to characterize and predict the future course of the pandemic in the West African countries. We estimated some specific characteristics of the disease’s dynamics such as its initial conditions, reproduction numbers, true peak and peak of the reported cases, with their corresponding times, final epidemic size and time-varying attack ratio. Our findings revealed a relatively low proportion of susceptible individuals in the region and in the different countries (1.2% across West Africa). The detection rate of the disease was also relatively low (0.9% for West Africa as a whole) and < 2% for most countries, except for Gambia (12.5 %), Cape-Verde (9.5%), Mauritania (5.9%) and Ghana (4.4%). The reproduction number varied between 1.15 (Burkina-Faso) and 4.45 (Niger), and the peak time of the pandemic was between June and July for most countries. Generally, the peak time of the reported cases came a week (7-8 days) after the true peak time. The model predicted 222,100 actual active cases in the region at the peak time, while the final epidemic size accounted for 0.6% of the West African population (2,526,700 individuals). The results obtained showed that COVID-19 has not severely affected West Africa as noticed in other regions of the world. However, current control measures and standard operating procedures should be maintained over time to accelerate a decline in the observed trends of the pandemic.

## Introduction

COVID-19 is a severe acute respiratory syndrome caused by the new coronavirus, SARS-CoV-2, which emerged from Wuhan, Hubei Province (China) towards the end of 2019 [1, 2]. It is currently, the most important threat to global public health. By August 15th, 2020, about 21,026,758 total confirmed cases and 755,786 deaths were recorded worldwide [3]. The disease has rapidly spread around the world (about 212 countries) [4] including the 54 African countries. By mid August 2020, The World Health Organisation (WHO) reported 936,062 and 152,483 confirmed cases and 18,286, and 2,351 deaths across Africa and the West-African region, respectively [3]. Healthcare services in the region have particularly faced critical times, making sensitive decisions regarding patients and their treatment [5]. It is clear that the COVID-19 pandemic has severely affected people’s life, health and economy. Actually, it led to an important increase in the demand of hospital beds and artificial respirators (mechanic and non-invasive). According to the WHO global health observatory data, most countries in West Africa have less than five hospital beds and two medical doctors per 10,000 of the population, while 50% of the countries have health expenditures lower than US$50 per capita [6]. In contrast, in European countries such as Italy and Spain, they have 34 and 35 hospital beds, respectively with 41 medical doctors, per 10,000 of the population, and US$2,840 and US$2,506 per capita expenditure [7]. Moreover, medical staff in the world have directly been exposed to infections [8]. Since vaccines are still under development, and antiviral drugs are not available for effective curative treatment of COVID-19 infections, the actual cure practice is hospitalization and intensive care unit management [9]. Prevention measures used are essentially non-pharmaceutical interventions such as regular hand washing with soap, mask wearing and social distancing. To be efficient, these non-pharmaceutical measures require a good understanding of the dynamic of the spread of the disease to aid in decision-making of their use.

Mathematical and statistical models can be useful tools to decisions making in public health. They are also important in ensuring optimal use of resources to reduce the morbidity and mortality associated with epidemics, through estimation and prediction [10]. The prediction of essential epidemiological parameters such as the peak time, the duration and the final size of the outbreak, is crucial and important for policy makers and the public health authorities to make appropriate decisions for the control of the pandemic [9]. Therefore, modelling and forecasting the numbers of confirmed and recovered COVID-19 cases could play an important role in designing better strategies for the control of the spread of COVID-19 in the world [4]. Since the appearance of the first COVID-19 case in the world, several studies have been conducted to model the dynamics of the disease. The main methods used were: deterministic modelling techniques [11–13], autoregressive time series models based on two-piece scale mixture normal distributions [4], stochastic modelling methods [14, 15], machine learning techniques [16,17], growth models [18,19] and bayesian method [17].

Among these modeling techniques, deterministic models are the most considered because of their simplicity. However, they fail to provide accurate results due to non-identifiability problem when the number of compartments and the number of parameters are high [2]. Actually, complex deterministic models have proven to be less reliable than simpler models such as the SIR model framework [2], which performs better in describing trends in epidemiological data. This under-performance may be worse when meta-population confirmed-cases data are considered. However, only few studies related to COVID-19 in Africa used mathematical models and prevalence data to study the dynamics, analyze the causes and key factors of the outbreak [1, 11, 20]. A recent study [11] assessed the current pattern of COVID-19 spread in West Africa using a deterministic compartmental SEIR-type model.

In this study, we used a simple deterministic susceptible–infectious–recovered (SIR)-type model to characterize and predict future trends of the spread of COVID-19 in West Africa. Specifically, we aimed to estimate some specific characteristics of COVID-19 dynamics (initial conditions of the pandemic, reproduction numbers, true peak, reported peak and their times and dates, final epidemic size and time-varying attack ratio). The originality of this work is that it focused on the 16 West African countries and the whole region as well. It is the first study dealing with the dynamics of the pandemic in each of the West African countries.

## Methods

### Model description

Problems of identifiability in parameters’ estimation in deterministic compartment models (especially complex models) are common in epidemiological modelling studies, which often imply biased estimations of parameters. In these situations, simpler models, which over-perform complex models in estimating reliable parameters, are recommended [2]. Hence, in this study, the SIR model [21] was considered with two removal rates as illustrated in the system below [22]:

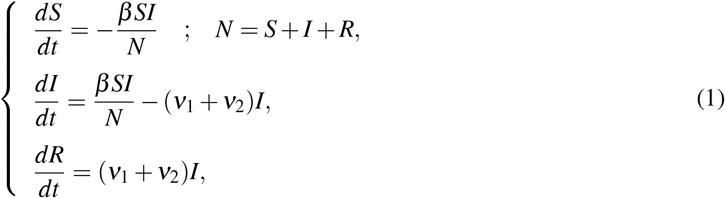

with initial conditions,

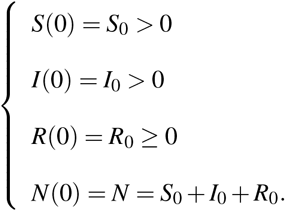

In Eq. (1), *S* = *S*(*t*), *I* = *I*(*t*) and *R* = *R*(*t*), representing the number of susceptible, infected and removed individuals at time t, respectively; while *N* is defined as the total population size for the disease transmission. The parameters *β*,*ν*_1_ and *ν*_2_ are the transmission rate, the removal rate of reported infected individuals (detected) and the removal rate of infected individuals due to all other unreported causes (mortality, recovery or other reasons), respectively. We considered the removal rate *ν*_2_ as constant with value *ν*_2_ = 1/10 [23]. From the second differential equation of Eq. (1), it could be noticed that *ν*_1_*I*_0_ represents the daily confirmed cases (*Ir*_0_) at time 0 of the outbreak. Thus, the relationship between the initial number of infected individuals and the detection rate, *ν*_1_, is *I*_0_=*Ir*_0_*Iν*_1_, was used in the estimation process.

### Data consideration and parameter estimation procedure

For each country, the data considered for the modelling spans the period from the date of detection of the first case of COVID-19 in the country and August, 12, 2020. Data considered were the daily numbers of reported cases that were assimilated to *ν*_1_*I*. These data were downloaded from the Global Rise of Education website [24]. Table 1 presents the demographic patterns [25], initial dates of the pandemic [24] and testing efforts (identification of new cases) of the countries [25]. We fitted the model (1) to the observed daily cases to study the dynamics of COVID-19 pandemic in the sixteen West African countries.

To improve the prediction power of Eq. (1), we used a cross-validation procedure of parameter estimation, where 90% of the observations were considered to estimate values of the five unknown model parameters (*S*_0_,*I*_0_,*R*_0_,*β*,*ν*_1_) and the remaining observations were used to validate the model. The Root Mean Square Error (RMSE) statistics was used as the measure of estimation precision:

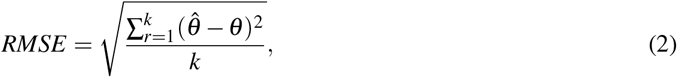

where 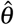 and *θ* are the predicted and observed number of daily cases respectively; *k* is the number of observations considered. We considered as *RMSE*_1_, the Root Mean Square Error computed on the 90% of the observations and *RMSE*_2_, computed on the remaining observations (10%)

The solutions of (1) were obtained using the built-in function *ODE45* of Matlab [26]. Then, the non linear least square estimate technique was performed to estimate the five parameters in (1) given starting values, using the built-in function *fminsearchbnd* of Matlab [26].

Afterwards, we simulated 2,000 different starting values of the five parameters using a resampling method (function *resample* of Matlab) for *S*_0_, *I*_0_ and *R*_0_ and the uniform distribution (function *rand* of Matlab) for *β* and *ν*_1_. Then, we estimated for each of the starting points, values of the five parameters using the non linear least square technique described above. The final values considered for these parameters were those related to both smallest values of *RMSE*_2_ and *RMSE*_1_ to guarantee a good fit of the model and a good predictive power. At the end of this process, we obtained reliable estimates of the model parameters with 95 % confidence intervals. Curves were plotted to show evolution trends of predicted daily new COVID-19, daily reported cases and the attack ratio for the 16 countries. With the estimated values of the five parameters from Eq. (1), the COVID-19 dynamic in each country was characterized by computing the following parameters with their 95% confidence intervals. Table 1 presents demographic patterns and testing efforts in the West African region.

**Table 1.** Demographic patterns and testing efforts of the 16 West African countries

| Countries | Population size | Density | First case | Tests/1M Pop | Cases/1000 tests |
| --- | --- | --- | --- | --- | --- |
| Benin | 12,123,200 | 108 | 03/16/2020 | 9,711 | 18 |
| Burkina-Faso | 20,946,992 | 76 | 03/10/2020 | - | - |
| Cape-verde | 556,498 | 138 | 03/21/2020 | 137,485 | 50 |
| Côte d'Ivoire | 26,428,999 | 83 | 03/12/2020 | 4,779 | 142 |
| Gambia | 2,421,823 | 239 | 03/17/2020 | 5,502 | 222 |
| Ghana | 31,072,945 | 137 | 03/12/2020 | 14,183 | 100 |
| Guinea | 13,160,021 | 53 | 03/14/2020 | 1,860 | 382 |
| Guinea-Bissau | 1,971,640 | 70 | 03/25/2020 | - | - |
| Liberia | 5,066,990 | 53 | 03/16/2020 | - | - |
| Mali | 20,294,900 | 17 | 03/26/2020 | 1,852 | 74 |
| Mauritania | 4,659,052 | 5 | 03/15/2020 | 14,980 | 100 |
| Niger | 24,269,389 | 19 | 03/22/2020 | 372 | 130 |
| Nigeria | 206,522,290 | 226 | 02/28/2020 | 1,943 | 134 |
| Senegal | 16,776,618 | 87 | 03/02/2020 | 8,632 | 93 |
| Sierra-Leone | 7,989,949 | 111 | 04/01/2020 | - | - |
| Togo | 8,293,924 | 152 | 03/07/2020 | 7,784 | 22 |
| West Africa | 402,555,230 | 66 | 02/28/2020 | - | - |

Date: 08/31/2020; Source: [25] and [24]; Tests/1M Pop: number of tests per 1 million individuals; Cases/1000 tests: number of positive cases per 1000 tests; - : no data.

-*Reproduction number*, 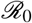 [27]: it is the average number of secondary infections, caused by an average infected individual (during his infectious period), in a fully constituted population:

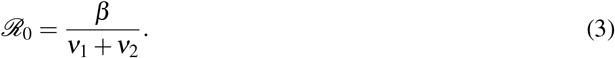

-*Running reproductive number*, 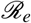 [27]: it measures the number of secondary infections caused by a single infected individual in the population at time t.

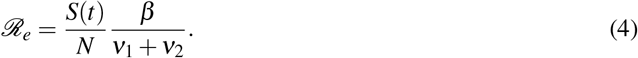

-*True peak size*, *n_pp_*, and *True peak time*, *T_pp_*. The true peak size indicates the largest daily number of new infectious cases in the population:

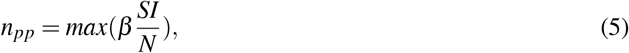

while the true peak time, *T_pp_*, represents the time at which the largest daily new infected cases is obtained. These two parameters were determined numerically.

-*Peak size of reported cases*, *n_rp_*, and *Peak time of reported cases*, *T_rp_*. The peak size of reported cases indicates the largest number of daily confirmed cases:

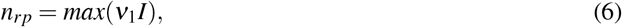

while the peak time of reported cases is the associated time to *n_rp_*. These were determined numerically.

-*Maximum number of active cases*, *I_max_*: since *I*_0_,*R*_0_ << *S*_0_, we assumed the number of initial susceptible individuals to be approximately equal to *N* (*S*_0_ ≈ *N*). Thus, *I_max_* can be approximated as follow [27]:

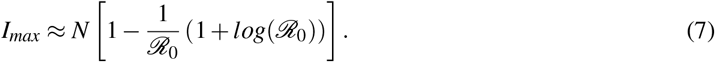

-*Final epidemic size*, *I_total_* [27]: it is the total number of cases over the course of the epidemic wave.

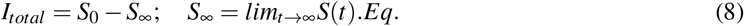

*S*_∞_ can be approximated considering the entire population as initially susceptible (*S*_0_ ≈ *N*); hence, following [27]:

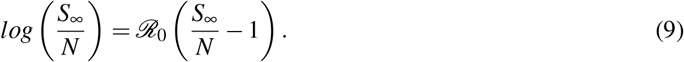

For each country, the equation Eq. (9) was solved numerically to determine *S*_∞_ through an iterative process.

-*Attack ratio*, *A_r_* [22]: it is the fraction of susceptible population that becomes infected. It is calculated along the epidemic wave as follows:

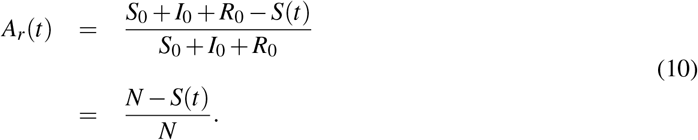

For West Africa as a whole, *S*_0_, *I*_0_ and *R*_0_ were first computed by summing the corresponding estimated values of the 16 countries. Afterwards, the model (9) was fitted to daily reported cases of the region using initial conditions computed. This allowed the estimation of *β* and *ν*_1_ and computation of the characteristics of COVID-19 dynamics across the region.

## Results

### Current patterns of COVID-19 transmission in West Africa

Table 1 reveals a great heterogeneity in the region in terms of population density and testing efforts. Countries like Cape-Verde, Mauritania and Ghana have put relatively more effort into identifying infected individuals while Niger, Nigeria and Guinea are countries with the lowest number of tests per 1 million people. Combining both the testing effort and the mean number of reported cases per test indicate a relatively less testing effort to identify many infected individuals (Guinea and Gambia). On the other hand, countries like Cape-Verde, Benin and Togo put much effort to find few COVID-19 cases (Table 1).

Results obtained from the estimation of initial conditions of COVID-19 pandemic in West Africa revealed the relatively low proportion of the susceptible individuals in most countries (about 1% of their total populations). However, countries such as Guinea-Bissau, Gambia and Cape-Verde showed relatively large proportion of susceptible individuals to COVID-19 with 17.5%, 6.0% and 2.4% of their total populations, respectively. The proportion of the susceptible individuals across West Africa was also relatively low (1.2%) (Tables 1 and 2). Moreover, before the detection of the first cases, infected individuals were present in the population of all the countries with some already recovered individuals. The detection rate of infected individuals was relatively low (less than 1%) for Benin, Burkina, Mali, Niger, Nigeria, Sierra-Leone and West Africa as a whole. However, some countries such as Gambia (12.5 %), Cape-Verde (9.5%), Mauritania (5.9%) and Ghana (4.4%) recorded the highest detection rates (Table 3).

In most countries, the model estimated an average of one new case of infection caused by an infected individual during the infectious period 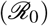, except for Sierra-Leone, Nigeria, and Côte d’Ivoire with 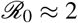 and Niger, which recorded the highest reproduction number 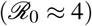 (Table 3).

## Long term dynamics of COVID-19 in West Africa

We analyzed the long term dynamics of COVID-19 in West Africa by first focusing on the true peak of the pandemic. In general, the estimated reported peak time came a week (7-8 days) after the true peak time in all the countries, while their estimated reported peak sizes accounted in average for 21% of the estimated true peak size (Table 3). Most countries had already experienced the peak of the epidemic wave. The true peak time was estimated in June for Sierra-Leone (14th), Mauritania (19th), Benin (27th), Côte d’Ivoire (24th) and Ghana (13th), while it was estimated in July for Liberia (4th) and Nigeria (5th), Senegal (18th), Guinea (23th), Cape-Verde (22th) and Gambia (28th). Niger recorded the earliest true peak time (April 8th), while the latest true peak time was on October 26th, 2020 for Burkina-Faso (Figure 1 and Table 3). The true peak time across the region was 1st July with 25,267 new cases (Table 3 and Figure 4a-b). Margin error of the estimated true peak time varied from one day to 16 days, with average value of five days in the region. Half of the countries (8 out of 16) recorded true peak size less than 1,000 new cases cases at the peak time. The highest numbers of new cases at the peak time were estimated at 19,021 and 17,703 for Niger and Nigeria, respectively (Table 3 and Figure 1). The estimates of the peak size of reported cases was 1,891 daily new cases across the region (Table 3 and Figure 4a-b).

**Table 2.** Estimates with 95% confidence intervals of the initial parameters of the SIR-model; *ν* _2_ = 0.10 is constant for all countries

| Country | $S_0$ ($\times 10^5$) | $I_0$ | $R_0$ | $\beta$ | $v_1$ | RMSE2 |
| --- | --- | --- | --- | --- | --- | --- |
| Benin | 1.212 [1.211-1.214] | 200 [197-203] | 39 [37-40] | 0.148 [0.145-0.151] | 0.005 [0.004-0.006] | 31.7 |
| Burkina-Faso | 2.175 [2.145-2.206] | 174 [164-183] | 160 [149-172] | 0.121 [0.121-0.122] | 0.006 [0.005-0.007] | 8.0 |
| Cape-Verde | 0.517 [0.460-0.573] | 21 [17-26] | 97 [91-102] | 0.226 [0.221-0.231] | 0.095 [0.092-0.098] | 98.8 |
| Côte d'Ivoire | 2.643 [1.265-4.021] | 73 [23-124] | 8 [6-9] | 0.178 [0.175-0.181] | 0.014 [0.011-0.017] | 65.6 |
| Gambia | 0.24 [0.21-0.26] | 8 [7-8] | 0 [0-2] | 0.259 [0.256-0.262] | 0.125 [0.124-0.126] | 15.2 |
| Ghana | 3.291 [2.388-4.193] | 46 [24-68] | 81 [80-83] | 0.20 [0.197-0.203] | 0.044 [0.041-0.047] | 462.0 |
| Guinea | 3.789 [3.428-4.151] | 229 [206-251] | 85 [82-88] | 0.146 [0.145-0.148] | 0.004 [0.004-0.005] | 35.7 |
| Guinea-Bissau | 3.458 [3.452-3.497] | 1990 [1970-2011] | 4 [2-5] | 0.146 [0.144-0.147] | 0.001 [0.000-0.004] | 15.2 |
| Liberia | 0.507 [0.506-0.508] | 100 [98-102] | 87 [85-89] | 0.146 [0.143-0.149] | 0.010 [0.007-0.013] | 153.4 |
| Mali | 2.240 [2.215-2.265] | 1175 [1139-1211] | 199 [194-203] | 0.167 [0.165-0.169] | 0.002 [0.000-0.005] | 39.1 |
| Mauritania | 0.466 [0.291-0.641] | 17 [8-26] | 42 [41-44] | 0.221 [0.216-0.225] | 0.059 [0.055-0.063] | 40.7 |
| Niger | 2.652 [1.578-3.276] | 1203 [1250-1296] | 39 [47-54] | 0.448 [0.441-0.455] | 0.001 [0.000-0.001] | 2.1 |
| Nigeria | 20.886 [10.651-31.121] | 252 [212-293] | 110 [102-117] | 0.173 [0.169-0.178] | 0.004 [0.000-0.009] | 2926.6 |
| Senegal | 1.976 [0.723-3.230] | 61 [16-138] | 99 [98-101] | 0.161 [0.160-0.163] | 0.016 [0.014-0.019] | 48.4 |
| Sierra-Leone | 0.799 [0.797-0.801] | 200 [191-209] | 5 [3-6] | 0.166 [0.160-0.171] | 0.005 [0.000-0.012] | 7.4 |
| Togo | 0.830 [0.662-0.999] | 42 [38-45] | 28 [26-29] | 0.144 [0.143-0.146] | 0.024 [0.022-0.026] | 8.0 |
| West Africa | 48.667 [31.820-62.722] | 5811 [5559-6194] | 1112 [1043-1144] | 0.152 [0.151-0.152] | 0.009 [0.007-0.010] | 1463.3 |

**Table 3.** Epidemiological statistics with their 95% confidence intervals indicating the dynamics of COVID-19 in the whole west African region and per country

| Country | $\mathcal{R}_0$ | $T_{rp}$ | $n_{rp}$ | $T_{pp}$ | $n_{pp}$ | $I_{max}(\times 10^4)$ | $I_{total}(\times 10^4)$ |
| --- | --- | --- | --- | --- | --- | --- | --- |
| Benin | 1.41 [1.38-1.43] | 114 [112-116] | 29 [24-34] | 104 [103-107] | 634 [558-710] | 0.57 [0.53-0.61] | 6.3 [6.3-6.4] |
| Burkina-Faso | 1.15 [1.14-1.15] | 239 [234-245] | 12 [11-13] | 229 [223-235] | 218 [213-222] | 0.19 [0.19-0.20] | 5.4 [5.3-5.4] |
| Cape-Verde | 1.16 [1.14-1.18] | 149 [148-150] | 50 [46-53] | 144 [143-145] | 103 [89-117] | 0.05 [0.04-0.06] | 1.4 [1.2-1.5] |
| Côte d'Ivoire | 1.56 [1.54-1.58] | 113 [112-114] | 269 [263-274] | 105 [104-106] | 2368 [1038-3698] | 1.97 [0.73-3.20] | 16.4 [7.2-25.6] |
| Gambia | 1.15 [1.15-1.16] | 138 [136-140] | 28 [29-30] | 134 [132-136] | 52 [51-53] | 0.02 [0.00-0.05] | 0.56 [0.42-0.62] |
| Ghana | 1.39 [1.38-1.40] | 131 [129-133] | 632 [629-634] | 124 [123-125] | 2138 [1413-2863] | 1.44 [0.76-2.13] | 16.6 [10.9-22.3] |
| Guinea | 1.40 [1.39-1.41] | 141 [139-143] | 76 [66-95] | 132 [130-134] | 1861 [1659-2063] | 1.71 [1.53-1.90] | 19.3 [18.2-20.5] |
| Guinea-Bissau | 1.44 [1.43-1.46] | 83 [82-84] | 20 [19-20] | 73 [72-74] | 2084 [2040-2128] | 1.85 [1.81-1.88] | 18.7 [18.6-18.9] |
| Liberia | 1.33 [1.32-1.33] | 120 [117-125] | 17 [17-18] | 111 [110-112] | 196 [193-198] | 0.17 [0.17-0.18] | 2.3 [2.2-2.3] |
| Mali | 1.64 [1.63-1.66] | 68 [64-72] | 35 [34-37] | 59 [55-63] | 2254 [2189-2320] | 2.00 [1.94-2.06] | 1.48 [1.47-1.49] |
| Mauritania | 1.39 [1.36-1.42] | 103 [101-104] | 119 [116-123] | 97 [95-99] | 331 [243-420] | 0.20 [0.05-2.18] | 2.3 [2.1-2.5] |
| Niger | 4.45 [4.37-4.52] | 20 [12-28] | 86 [84-87] | 15 [6-24] | 19021 [18597-19444] | 10.72 [10.49-10.96] | 26.2 [20.0-27.9] |
| Nigeria | 1.67 [1.64-1.69] | 121 [117-125] | 774 [608-824] | 112 [108-116] | 21800 [6616-36983] | 19.50 [11.89-27.11] | 141.0 [126.7-249.8] |
| Senegal | 1.38 [1.37-1.39] | 147 [146-152] | 136 [133-138] | 139 [138-140] | 989 [55-2033] | 0.82 [0.02-1.81] | 9.7 [1.8-17.7] |
| Sierra-Leone | 1.58 [1.56-1.60] | 83 [78-88] | 31 [30-33] | 75 [69-81] | 700 [673-727] | 0.62 [0.59-0.64] | 5.0 [4.9-5.2] |
| Togo | 1.16 [1.16-1.17] | 220 [218-222] | 21 [15-27] | 212 [210-214] | 110 [100-121] | 0.09 [0.08-0.09] | 2.2 [2.0-2.4] |
| West Africa | 1.39 [1.38-1.41] | 119 [117-121] | 1891 [1874-1909] | 111 [108-112] | 25267 [24239-26294] | 22.21 [21.21-23.22] | 248.8 [140.8-250.7] |

The final epidemic size accounted for 0.6% of the population of West Africa. This estimate was generally low (< 1% of the population size) for more than half of the countries, though Guinea-Bissau and Cape-Verde recorded the highest final epidemic sizes in terms of proportion to their population size (> 9%). The estimates of the maximum number of actual daily active cases at the peak time for most countries were greater than 1,000 cases, though, it was 107, 200 and 195, 000 for Niger and Nigeria, respectively (Table 3).

**Fig 1.**
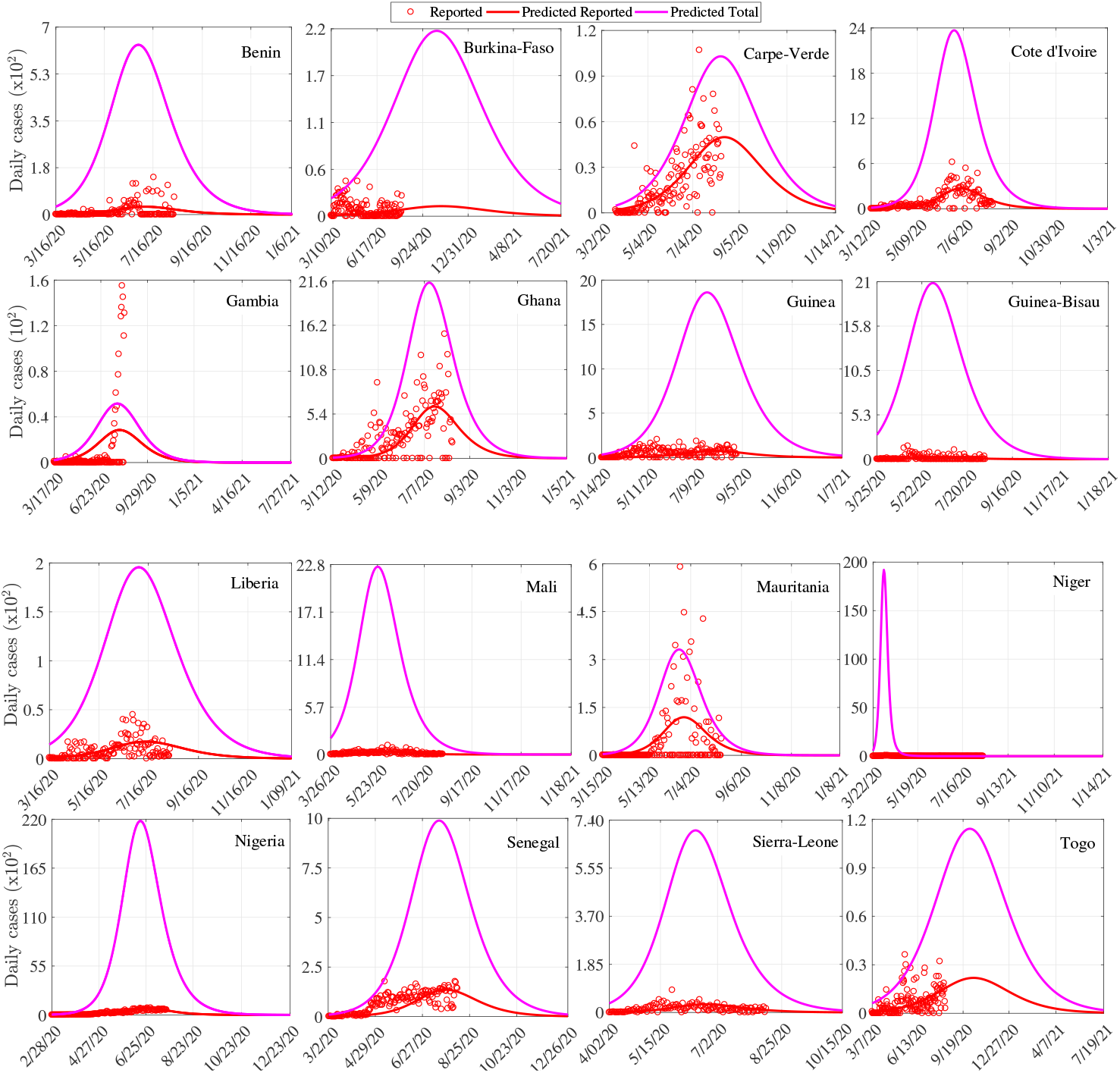
Evolution trend of the COVID-19 daily cases per country.

The running reproduction number helped to assess the evolution trends of the disease. It decreased over time in all the countries from the beginning of the outbreak (1.2 −4.5) to a stable point, which varied according to countries (0.50 − 0.82; Figure 2). As expected, the fraction of susceptible individuals being infected (attack ratio) increased over time from 0% to 40% − 70%, depending on countries. These evolving trends in the reproduction number and attack ratio were similar to those noted for West Africa as a whole (Figure 4b-c).

**Fig 2.**
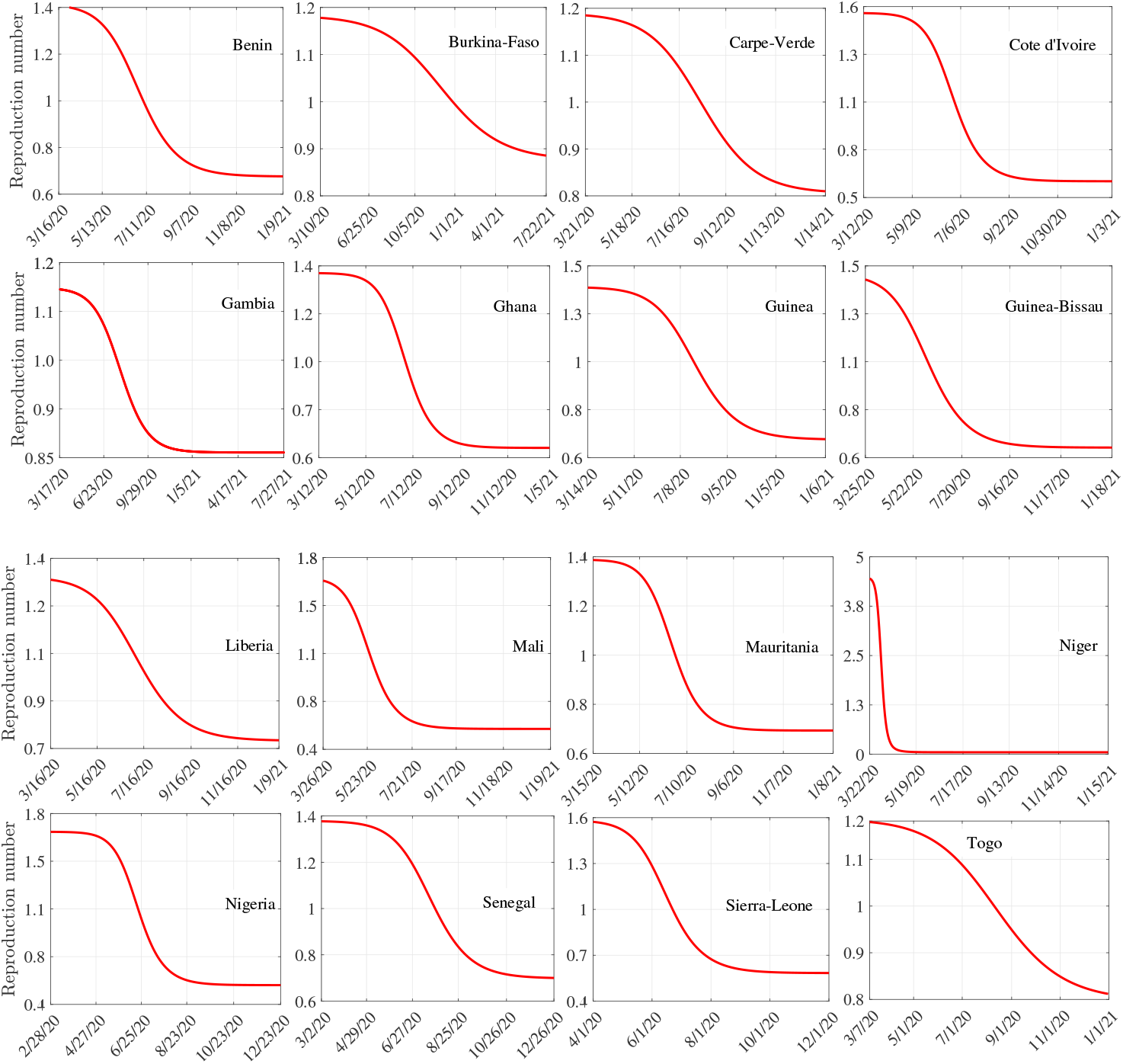
Running reproduction number per country in West Africa.

**Fig 3.**
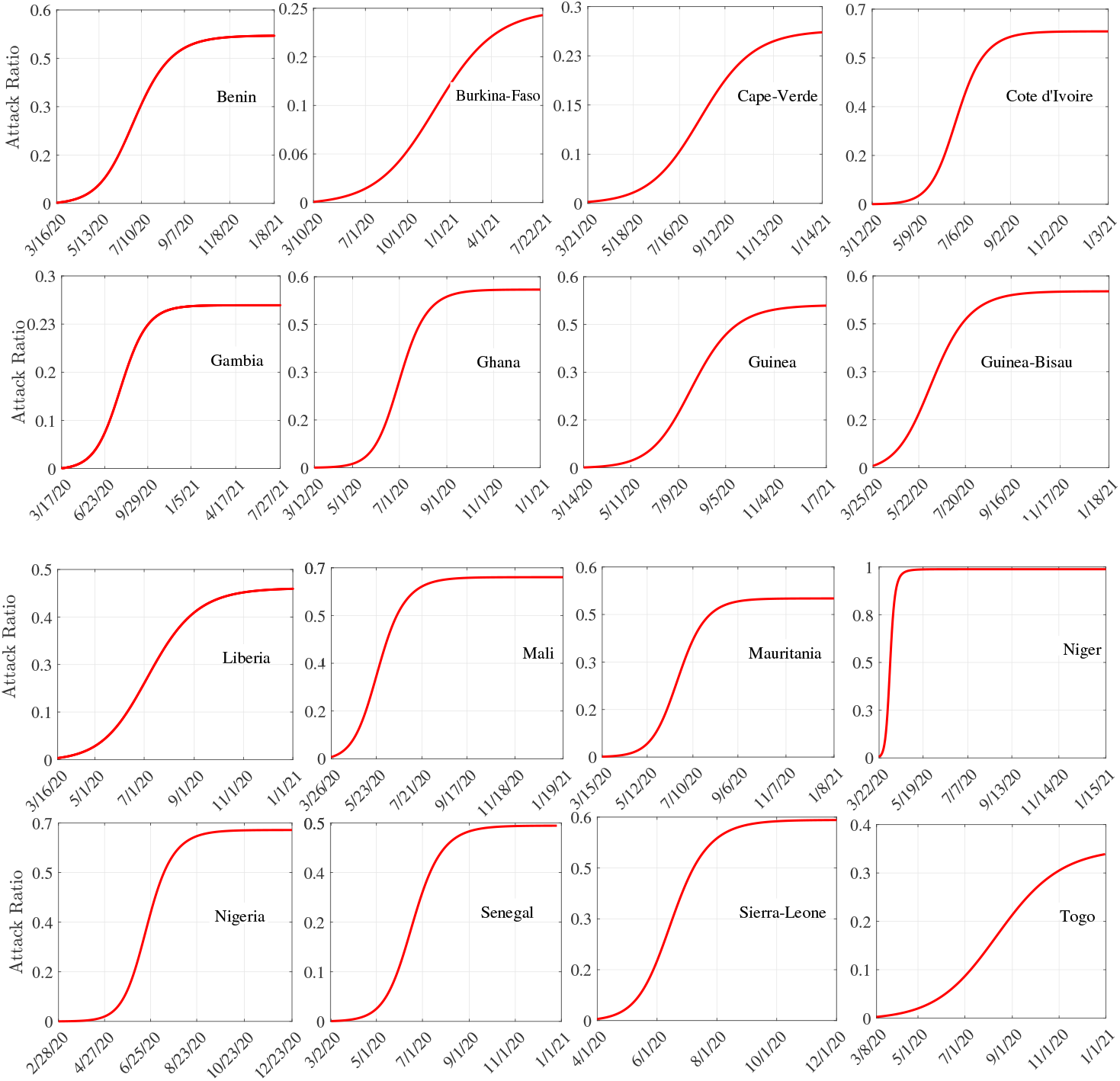
Evolution trend of the attack ratio per country in West Africa.

**Fig 4.**
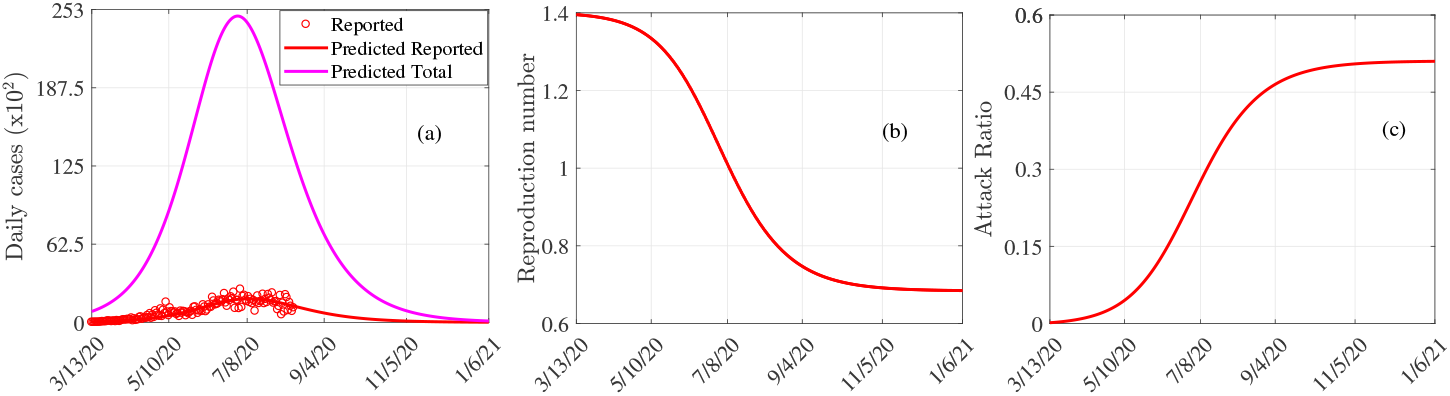
Trends in COVID-19 dynamics across West Africa. (a) Prediction of the true peak and reported peak date, and size of COVID-19. (b). Evolution trends of the reproduction number. (c) Evolution trends of the attack ratio.

## Discussion

In epidemiology, understanding the dynamics of an epidemic outbreak and predicting its future course is a major research question, which is often studied using modelling techniques [28, 29]. Estimation and prediction rely on mathematical and statistical models, which inform public health decision makers and ensure optimal use of resources to reduce the morbidity and mortality associated with epidemics [10, 30–32]. For instance, estimation of epidemiological parameters and prediction on the *Influenza* outbreak dynamics in Canada was done using the Richard’s model [30], while a three-parameter logistic growth model was used to study and forecast the final epidemic size in real-time of the *Zika* virus outbreaks in Brazil from 2015 to 2016 [28].

In this study, we used a deterministic SIR-type model to understand COVID-19 dynamics in the West African countries and estimated the overall number of susceptible individuals, which accounted for the 1.2% of the West African population and 1% for most countries, except Guinea-Bissau and Cape Verde, where the susceptible individuals accounted for more than 2%. The concept of susceptibility in this study, considering the model (1) takes into account sensibility and exposure to COVID-19. Thus, the low proportion of susceptible individuals in most countries could be explained by physiological factors, immunity acquired from other diseases or epidemics, or low levels of exposure to the disease (majority of individuals are far from the epicenter of the pandemic in different countries, with relatively low human movement between remote regions in West Africa as compared to the developed countries). Actually, individuals who are more susceptible or more exposed tend to be infected earlier, depleting the susceptible subpopulation of those who are at higher risk of infection [33]. This selective depletion of susceptible individuals intensifies the deceleration in the incidence. Our findings also revealed a great disparity between countries in terms of the testing rates of COVID-19. Countries such as Guinea and Gambia and to a less extent, Côte d’Ivoire and Nigeria, showed a relatively less testing effort to identify many infected individuals. This suggests that there may not be enough tests being carried out to properly monitor the outbreak [24]. In contrast, countries such as Cape-Verde, Benin and Togo, which have recorded less than or equal to 50 positive cases per 1,000 tests, seem to be effectively controlling the pandemic according to the WHO criteria [34]. Compared to relatively wealthier countries such as Australia, South Korea and Uruguay, it takes hundreds of tests to find one case [24].

The detection rate considered in model Eq. (1) is a better indicator of the testing effort, since it represents the proportion of active cases in the population that are identified daily. Our results revealed a relatively low detection rate of COVID-19 in West Africa with less than 2% in most countries except Gambia, Cape-Verde and Mauritania (> 5%). The last two countries are also the ones with the highest testing rates (table 1) confirming the link between detection rate and testing effort [35]. The fact that Gambia had a relatively low testing effort and a high detection rate suggests that either an efficiency in the contact tracing and testing efforts directed towards those really exposed, or a poor quality of parameter estimation. Indeed, especially for Gambia, the peak size of the reported cases was underestimated by the model compared to observed data. This may be explained by an unusually long period of very low detection of the disease in the country (March 18 -July 13, 2020). Other factors that may explain significant differences between some estimates and observed data, including data quality are discussed elsewhere [36]. For example, data from Benin showed, from June 30, long periods (4-7 days) without detected cases, often alternating with a large number of detected cases (more than 80) within a single day. This may indicate a possible problem in the daily reporting of data or in the planning of PCR tests and has led to a significant underestimation of the peak of reported cases (Table 3).

The fairly low detection rates in most West African countries demonstrate low testing effort and may be explained by a number of factors, including the availability of testing kits and qualified healthcare workers as well as low ability to control the disease due to their low GDP. For instance, the average detection rate of COVID-19 in the world in April 2020 was estimated at 6% [37]. It is also useful to note that the estimated average detection rate hides a great variability in the testing effort over time. Indeed, it is generally accepted that the testing rate is relatively low at the very beginning of an epidemic outbreak, but can increase rapidly over time when better response mechanisms are put in place [38].

The dynamics of COVID-19 in West Africa shows a reproduction number greater than one in all countries (from 1.2 in Burkina-Faso to 4.4 in Niger), while it was 1.4 for the whole region. This value is relatively low compared to 1.6, estimated for the same region in June in a recent study using a modified SEIR model [11]. Hence, this reveals either a declining trend in the pandemic over time, or the result of using a different modeling approach to estimate the reproduction number. COVID-19 appears more serious than the Ebola outbreak in Africa given the reproduction number. Indeed, the reproduction number was estimated at 1.1, 1.2 and 1.2 for Guinea, Liberia and Sierra-Leone, respectively, against 1.4, 1.3 and 1.6 for the same countries as far as COVID-19 is concerned [39]. These comparisons indicate that COVID-19 is on average 1.29 times more reproducible than Ebola in these countries.

The trend in the running reproduction number reveals a rapid decline in all the countries except Burkina-Faso, and reveals some efficiency of the control measures put in place and being implemented in the different countries. Most countries had already experienced the peak in the new COVID-19 cases in June and July. In general, the reported peak was very low compared with the true peak.

However, current control measures should be maintained overtime to avoid a possible second wave as observed in other parts of the world.

## Conclusion

Our study shows that the novel COVID-19 pandemic, although highly contagious has not seriously impacted West Africa in terms of prevalence, compared to other parts of the world, particularly Europe and the USA. Actually, the total number of susceptible individuals and final epidemic size account for 1.2% and 0.6% of the total population size of West Africa, respectively. But, the relatively low reported cases are related to very low testing effort in the West African countries. The study also indicates a relatively low detection rate, and for most countries in the region, the dates of the true peak of infection have passed (June-July 2020). Nevertheless, the pandemic is still ongoing in the region and it is important that the non-pharmaceutical measures currently in place should continue over time to help reduce its spread dynamics, pending adequate effective treatment.

## Data Availability

I declare that all data referred in the manuscript are available.

https://ourworldindata.org/coronavirus-source-data

## Acknowledgments

HBT acknowledges the support of IMU-CDC through GRAID program. RGK acknowledges the support from the African German Network of Excellence in Sciences (AGNES) and the Alexander von Humboldt Foundation (AvH).

## Authors’ contributions

RGK conceived the ideas, designed the methodology, wrote the codes and supervised the work; SHH gathered the data, run the codes for some countries and wrote most sections of the manuscript; HBT run the codes for some countries and write some sections of the manuscript. All authors reviewed the drafts and gave final approval for publication.

